# Endothelial sampling *in situ* enables genetic characterization of vein of Galen Malformation

**DOI:** 10.1101/2023.12.01.23299300

**Authors:** Andrew T. Hale, Shanrun Liu, Fengyuan Huang, Yuwei Song, Michael R. Crowley, David K. Crossman, Caroline Caudill, Anastasia A. Smith, Lindsey Chapman, Michael J. Feldman, Benjamin W. Saccomano, Brandon G. Rocque, Curtis J. Rozzelle, Jeffrey P. Blount, James M. Johnston, Zechen Chong, Jesse G. Jones

**Affiliations:** Department of Neurosurgery, University of Alabama at Birmingham, Birmingham, AL; Heflin Genetics Center and Genetics Research Division, University of Alabama at Birmingham, Birmingham, AL; Single Cell and Flow Cytometry Core, University of Alabama at Birmingham, Birmingham, AL; Division of Pediatric Neurosurgery, Children’s of Alabama, Birmingham, AL

**Keywords:** vein of Galen malformation, vein of Galen aneurysmal malformation, genomics, human genetics, functional genomics, endoluminal biopsy, endothelial biopsy, endovascular biopsy, somatic mutation, single cell RNA sequencing

## Abstract

**Background and Objectives:** Vein of Galen malformation (VOGM), the result of arteriovenous shunting between choroidal and/or subependymal arteries and the embryologic prosencephalic vein, is among the most severe cerebrovascular disorders of childhood. While endovascular treatment options have improved outcomes, morbidity and mortality remain high. We hypothesized that *in situ* analysis of the VOGM lesion using endoluminal tissue sampling (ETS) is feasible and may advance our understanding of VOGM genetics, pathogenesis, and maintenance.

**Methods:** We collected germline DNA (cheek swab) from patients and their families for genetic analysis. *In situ* VOGM ‘endothelial’ cells (EC), defined as CD31+ and CD45-, were obtained from coils via ETS during routine endovascular treatment. Autologous peripheral femoral ECs were also collected from the access sheath. Single-cell RNA sequencing (scRNA-seq) of both VOGM and peripheral ECs was performed to demonstrate feasibility to define the transcriptional architecture. Comparison was also made to a published normative cerebrovascular transcriptome atlas. A subset of VOGM ECs was reserved for future DNA sequencing to assess for somatic and second-hit mutations.

**Results:** Our cohort contains 6 patients who underwent 10 ETS procedures from arterial and/or venous access during routine VOGM treatment (aged 12 days to ∼6 years). No periprocedural complications attributable to ETS occurred. Six unique coil types were used. ETS captured 98 ± 88 (mean ± SD; range 17-256) experimental ‘endothelial’ cells (CD31+ and CD45-). There was no discernable correlation between cell yield and coil type or route of access. Single cell RNA sequencing demonstrated hierarchical clustering and unique cell populations within the VOGM EC compartment compared to peripheral EC controls when annotated using a publicly available cerebrovascular cell atlas.

**Conclusion:** ETS appears safe and may supplement investigations aimed at development of a molecular-genetic taxonomic classification scheme for VOGM. Moreover, results may eventually inform the selection of personalized pharmacologic or genetic therapies for VOGM and cerebrovascular disorders more broadly.

## Introduction

Vein of Galen malformation (VOGM) is a congenital high flow vascular lesion characterized by aberrant communication between the choroidal or subependymal cerebral arteries and median prosencephalic vein of Markowski. Arteriovenous (AV) shunting can lead to congestive heart failure in neonates and hydrovenous disorder of early childhood.^1^ The latter condition involves venous hypertension with consequent brain atrophy, hydrocephalus, and epilepsy. Current methods for closing the AV shunt rely on percutaneous trans-arterial or trans-venous access with microcatheters, followed by vascular occlusion with liquid embolics and/or coils.^2^ Endovascular embolization represents the primary treatment but typically requires several procedures, staged over months to years. Sequelae of prolonged venous hypertension during this period lead to permanent disability or death in ∼40% of patients.^2,3^ Intervention in the neonatal stage also carries a significant risk of peri-procedural stroke and death.^4^ Thus, safer and more efficacious VOGM therapies are urgently needed.

Little is known about how VOGM develops between the 6th and 11th weeks of gestation^5^ or how it maintains itself as a large vascular network consuming up to 70% of cardiac output.^6^ These fundamental knowledge gaps impede development of improved therapies. Human genetics and functional genomics are well suited to VOGM because they integrate static (either inherited or somatic mutation) and dynamic (i.e., gene expression) approaches. For a congenital disease that lacks robust animal models, genomic analysis will complement observational and experimental studies. Specifically, whole-exome sequencing (WES) represents an established method for identifying inherited, *de novo*, and copy number aberrations in disease, including VOGM.^7,8^ Functional genomics approaches such as single-cell RNA sequencing (scRNA seq) delineate molecular mechanisms underlying diseases with a genetic or shared molecular component.^9-16^ Collectively, these approaches may enable (1) detection of potential causative or risk-conferring mutations; (2) elucidation of cellular composition *in situ*; and (3) identification of convergent molecular mechanisms amenable to pharmacologic or even gene therapy based approaches.

Recently, an endothelial tissue sampling (ETS) technique has emerged for obtaining lesional tissue *in situ*.^17,18^ During routine endovascular treatment, a catheter is positioned within the pathologic region and a standard endovascular coil advanced into the vasculature. After ∼ 2 minutes contact with the vessel wall, it is completely withdrawn from the body ^19,20^ Adherent endothelial cells are subsequently isolated using antibody mediated selection and flow cytometry. ETS has facilitated rapid advancement in the understanding of brain aneurysm and arteriovenous malformation (AVM) genomics and proteomics.^21,22^ However, no study to our knowledge has reported successful use of ETS in patients with VOGM. Advances in genomic and molecular analyses now position ETS to enable discovery of mechanisms underlying VOGM pathogenesis and maintenance with the goal of developing personalized, biologically based therapies.

## Methods

### Study population

IRB approval was granted by the author’s institution for this prospective observational study. Parents provided informed consent. Six pediatric (range: 12 days to ∼6 years) VOGM patients undergoing standard of care embolization were recruited between 2022-2023. Diagnosis was confirmed by catheter angiography. Clinical presentation included neonatal congestive heart failure and hydrovenous disorder. All patients were seemed suitable for clinical intervention based on Bicetre score.^23^ During each endovascular procedure, ETS was performed if deemed safe by the primary operator (J.G.J.). Staged embolization patients underwent ETS on more than one occasion. Information regarding the sampling location, coil type and other data were recorded on a templated form and entered into a secured Redcap database. Germline DNA was collected from all subjects and first-degree relatives who were available for consent via cheek swab. Pedigrees were assembled to eventually compare germline DNA against EC’s retrieved by ETS to identify somatic and second-hit mutations.

### Endoluminal tissue sampling (ETS) procedure

ETS protocols have been established previously for other cerebrovascular lesions.^24-27^ However, VOGM poses novel challenges due to the shearing effect of high flowing blood and small patient size, which precludes many common endovascular devices. Clinical factors alone determined vessel selection, catheter selection, and access route. Trans-arterial embolization more often characterized earlier stages of treatment. Flow directed Magic (Balt, Montmorency) microcatheters are not amenable to ETS. Thus, the sample selection was limited to treatments employing a Marathon microcatheter (EV3, Irvine CA) which accommodates smaller (up 6 mm) Blockade coils. Achieving a stable coil nest in such high flow pedicles is challenging and best done at a curve or narrowing in the vessel. Trans-venous embolization often allowed standard 0.017” microcatheters (and larger coils) given the less tortuous and delicate access route, except in cases of retrograde pedicle catheterization when an Apollo (EV3, Irvine CA) was used in conjunction with Onyx. Like Marathon, Apollo accommodates smaller Blockade coils for ETS. Coils were sized to approximate the vessel’s diameter and conform to its walls. After 2 minutes of endoluminal dwell time, the coil was resheathed through the microcatheter, protecting it from contact with other vessel segments. At procedure’s end, the vascular access sheath(s) were collected, and ECs were isolated.

### Cell isolation and dissociation from coils

The explanted coil is immediately placed into a conical tube of DMEM on ice. The coil is then transported to the flow cytometry core within 30 minutes. DMEM is aspirated and the coil rinsed in DPBS (without Ca or Mg), incubated in 0.25% Trypsin-EDTA at room temperature for 5 minutes prior to neutralization with 1ml of FBS. The coil is then washed with a trypsin-FBS solution to ensure complete dissociation. The dissociate is then spun down (∼500g, 5 minutes), the supernatant removed, and the pellet resuspended in ACK lysis buffer on ice, for 3∼4 mins to lyse red blood cell contaminants. The ACK lysis buffer is then neutralized using 1X PBS or DMEM. Cells are pelleted again and resuspended in PBS 100ul with Alexa Fluor 647–conjugated monoclonal anti-human CD31 antibody (BD Biosciences, San Jose, CA; 1:200 dilution) and anti-human CD45 antibody (BD Biosciences, San Jose, CA; 1:200 dilution). The cell solution containing CD31 and CD45 antibodies are incubated on ice for 20 minutes prior to addition of propidium iodide (to exclude dead cells, 120ug/ml, 1:100 dilution in DPBS without Ca or Mg) for an additional 10-minute incubation on ice. Finally, 500ul DPBS (without Ca or Mg) is added, centrifuged for 5 minutes at 500g, the supernatant is removed, the cell pellet is resuspended in 300ul DPBS, and individual cells are sorted into a 96-well plate. Cells obtained from the shealth are processed in an analogous fashion.

### Cell sorting and flow cytometry

Cells are subsequently fluorescently sorted on a BD FACSAria II Flow Cytometer (BD Biosciences). Nonviable cells were excluded based on propidium iodide (PI) positivity, monocyte populations were excluded based on CD45 positivity, and endothelial enrichment was performed through positive selection of CD31 cells. Viable cells enriched for endothelium were therefore considered to be CD31-positive, CD45-negative, and PI-negative cells. Cells are then sorted into single-cells in a 96-well plate (already containing 4ul of DPBS as indicated above) for either single-cell or bulk analysis. The plate is then sealed and frozen at -80°C for downstream human genetic and functional genomic analyses.

### Single cell RNA sequencing

Raw sequencing data were processed with the Cell Ranger pipeline software (v.3.0.2; 10x Genomics). The Cell Ranger count pipeline was used to perform quality control, sample demultiplexing, barcode processing, alignment, and single-cell 5’ gene counting. Cell ranger “count” was used to align raw reads against the hg38 genome (refdata-gex-GRCh38-2020-A) using CellRanger software (v.4.0.0) (10x Genomics). Subsequently, cell barcodes and unique molecular identifiers (UMI) underwent filtering and correction using default parameters in Cell Ranger. Reads with the retained barcodes were quantified and used to build the gene expression matrix.

Seurat (v.3.0.0), implemented using the R package, was applied to exclude low-quality cells.^28^ Cells that expressed fewer than 200 and larger than 5000 genes were filtered out. The processed data was normalized using Seurat’s ‘NormalizeData’ function, which used a global scaling normalization method, LogNormalize, to normalize the gene expression measurements for each cell to the total gene expression. Highly variable genes were then identified using the function ‘FindVariableGenes’ in Seurat. The anchors were identified using the ‘FindIntegrationAnchors’ function, and thus the matrices from different samples were integrated with the ‘IntegrateData’ function. The variation arising from library size and percentage of mitochondrial genes was regressed out using the function ‘ScaleData’ in Seurat. Principal component (PC) analysis was performed using the Seurat function ‘RunPCA’, and K-nearest neighbor graph was constructed using ‘FindNeighbors’ function in Seurat with the number of significant PCs identified from PCA analysis. Clusters were identified using ‘FindClusters’ function with resolution of 0.4. The clusters were visualized in two dimensions with UMAP. The normalization, integration, and clustering were performed under standard Seurat workflow.

## Results

All six families that were solicited to participate in the study agreed and provided informed consent. ETS was performed a total of ten times (owing to staged procedures involving the same subject), in three arteries and seven veins. All coils yielded CD31+, CD45-experimental cells, although the number varied widely (98 ± 88 mean ± SD; range 17-256) (**Table 1**). Yield tended to improve later in the course of treatment (for subjects who underwent >1 ETS), suggesting flow reduction may shear fewer cells off the coil. No periprocedural complications related to ETS were encountered and no subjects were lost to follow-up.

**Table 1.** Overview of ETS results from patients with VOGM indicating participant ID, age at ETS, ethnicity, route of access for routine treatment and ETS, vessel sampling, coil type used for ETS, and experimental cell (i.e., CD31+, CD45-) yield. PVM = prosencephalic vein of Markowski. No patient identifiers were available to anyone outside of the research group. Age ranges were determined as follows: neonates (< 1 month), infant (1 month ≤ 2 years), and children (≥ 2 years).

|  | Age range at ETS | Ethnicity | Access route | Vessel | Coil type | Experimental cell yield (#) |
| --- | --- | --- | --- | --- | --- | --- |
| VOGMUAB_001 | Child | Black | Arterial | Subependymal branch of L P1 | Balt Barricade (1x2mm) | 256 |
| VOGMUAB_001 | Child | Black | Venous | Ampulla | Cerenovous (3x6mm) | 217 |
| VOGMUAB_002 | Child | White | Venous | PVM | Cerenovus Deltafill (8x35mm) | 20 |
| VOGMUAB_002 | Child | White | Venous | PVM | Cerenovus Deltafill (8x35mm) | 80 |
| VOGMUAB_003 | Child | Mixed | Venous | PVM | Balt Barricade (6mm x 20cm) | 17 |
| VOGMUAB_005 | Infant | Black | Arterial | R posterior choroidal artery | Balt Barricade (1mm x 3cm) | 24 |
| VOGMUAB_006 | Child | Black | Venous | PVM | Cerenovus Galaxy G3 12 (12mm x 30cm) | 178 |
| VOGMUAB_010 | Neonate | White | Arterial | L posterior choroidal artery | Balt Barricade (4mm x 8cm) | 18 |
| VOGMUAB_010 | Neonate | White | Venous | Ampulla | Balt Barricade (4mm x 6cm) | 96 |
| VOGMUAB_010 | Neonate | White | Venous | Ampulla | Balt Barricade (5mm x 10cm) | 78 |

This report focuses on technical aspects of ETS in VOGM. Complete trios were assembled in 2/6 families, mother and child only in 3/6, and father and child only in 1/6 (**Figure 1**). Cheek swab DNA from all individuals listed in **Figure 1** was collected. An overview of our approach can be found in **Figure 2**. ETS is performed (see Methods for full details) and specimens are immediately transported for flow cytometry sorting. After positive selection with CD31 and negative selection of CD45, experimental ‘endothelial’ cells are obtained (**Figure 3**). Pooled experimental cells are then lysed for DNA purification using standard methodologies.^29^ This finding demonstrates ETS’ potential to enable identification of coding mutations, copy number variants, deletions, insertions, genomic rearrangements, and the single cell/nucleus RNA transcriptional landscape unique to VOGM tissue *in situ*. Finally, single cell RNA sequencing (scRNA seq) was performed to demonstrate feasibility of this approach on VOGM samples obtained through ETS (**Figure 4**). Using hierarchical clustering, we identify 12 unique cell populations between VOGM *in situ* and femoral access samples. Collectively, these data illustrate the feasibility of ETS for VOGM and we offer an integrative strategy for advancing our molecular genetic understanding of VOGM pathogenesis and maintenance.

**Figure 1.**
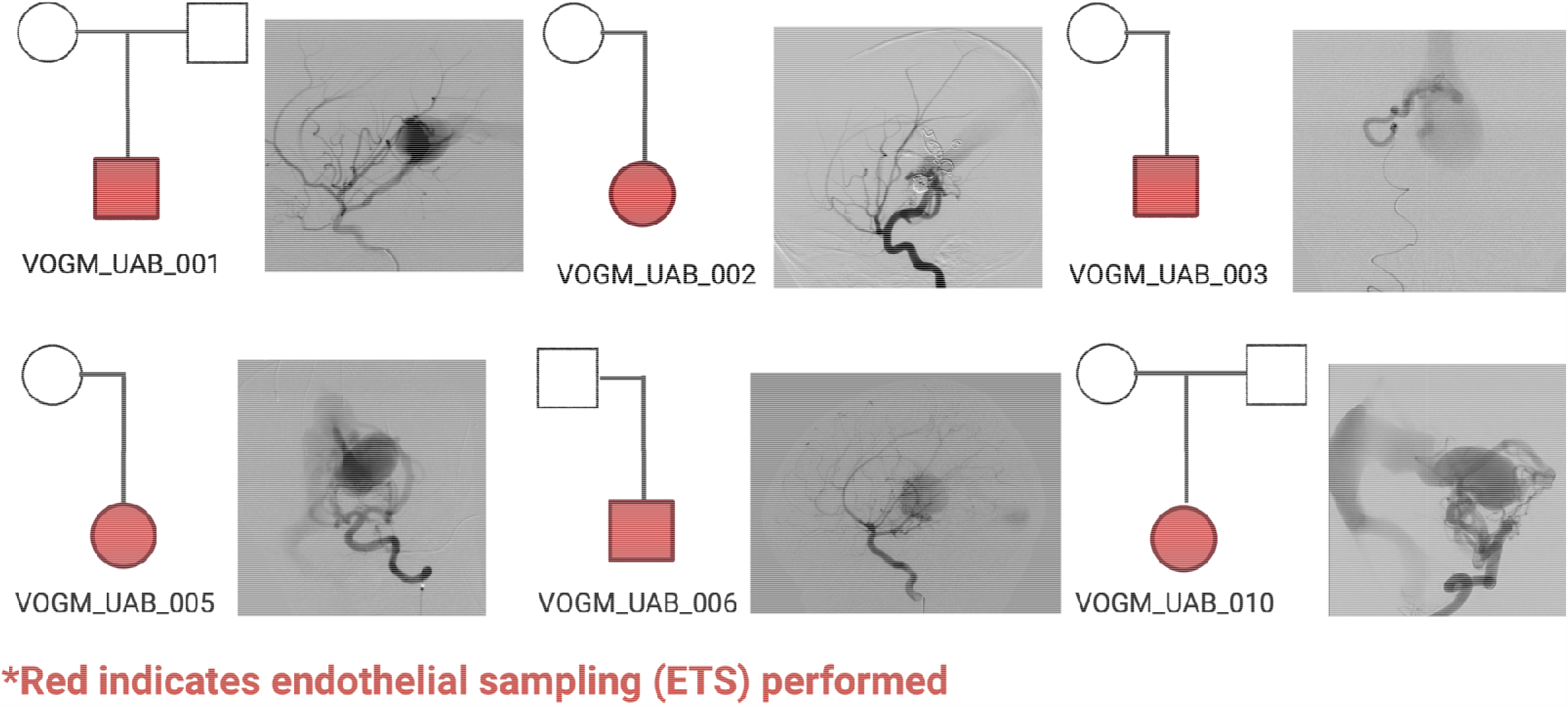
Pedigrees of VOGM families where the VOGM patient (proband) underwent endothelial sampling (red). Males are indicated with squares and females indicated with circles. No patient identifiers were available to anyone outside of the research group.

**Figure 2.**
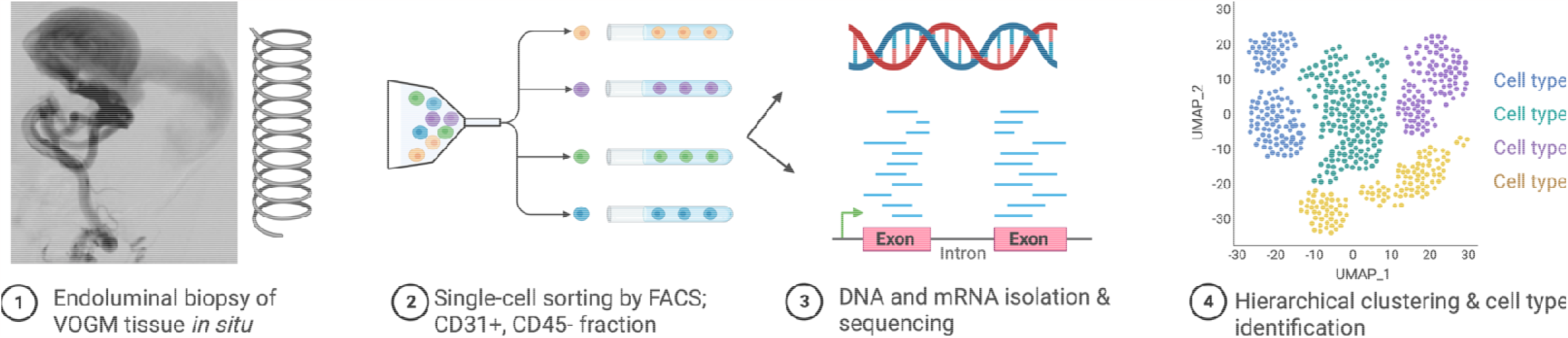
Overview of our approach to elucidate the genetic basis of VOGM. Figure was designed using Biorender.

**Figure 3.**
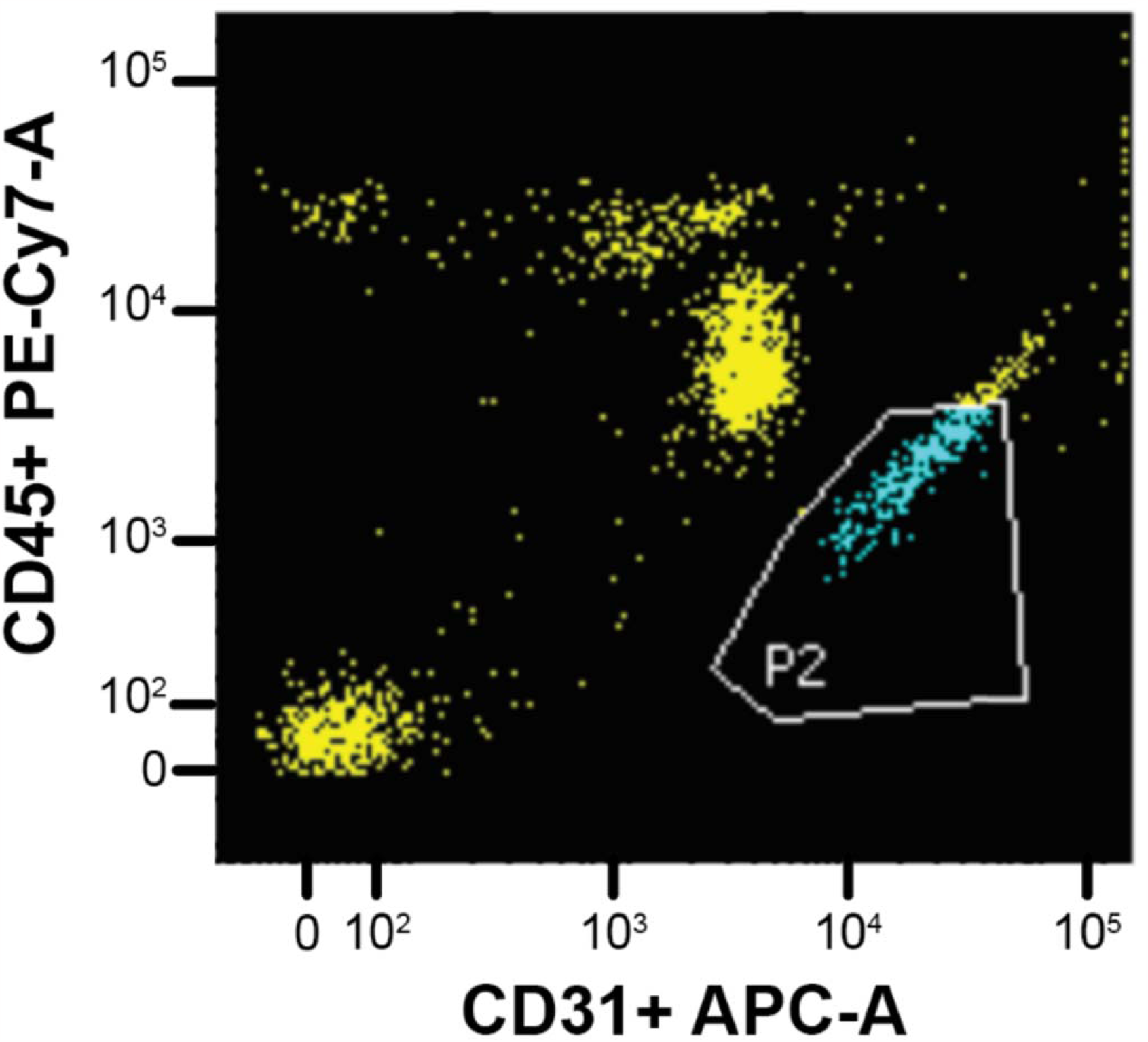
Representative flow cytometry plot demonstrating experimental ‘endothelial’ cell population (blue; CD31+, CD45-) obtained during ETS of VOGM lesion. The yellow cells represent the excluded fraction which are likely monocyte and red blood cell in origin.

**Figure 4.**
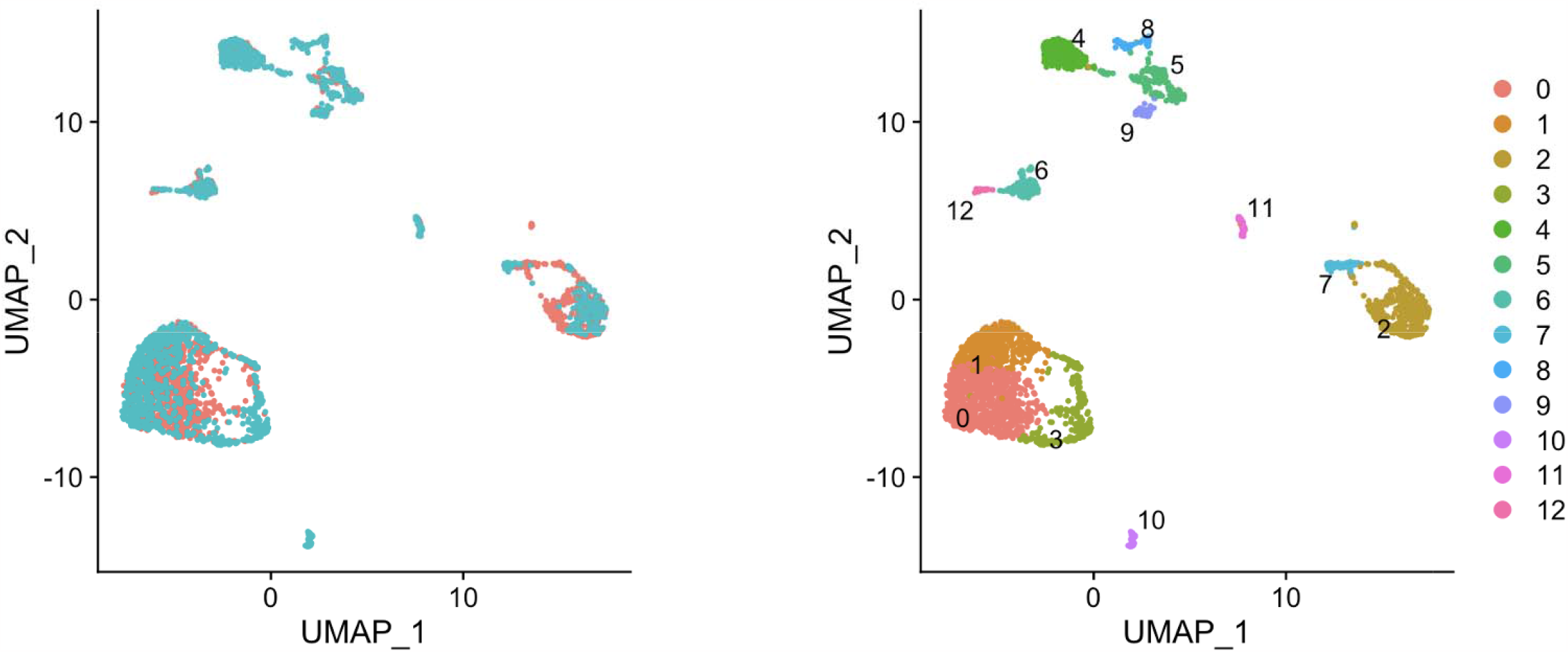
Single cell RNA sequencing (scRNA) of VOGM *in situ* identifies distinct cell types. (A) Pink represents ECs (CD31+, CD45 -) obtained from the femoral access sheath and blue represents *in situ* VOGM ECs obtained by ETS. (B) Hierarchical clustering identifies 12 unique cell populations across control and VOGM ECs using a published cerebrovascular transcriptome atlas for annotation.^30^

## Discussion

While VOGM represents a severe cerebrovascular anomaly with profound systemic and neurologic consequences,^30^ it is also rare and until now, pathologic studies were limited to post-mortem specimens.^31^ For these reasons, investigative studies into VOGM embryologic development, vascular maintenance, post-natal growth through angiogenesis, and cellular response to treatment have been hindered. Recent WES studies have identified putatively causative *de novo* and inherited gene mutations in ∼30% of sporadic VOGM cases.^8,32^ These studies identified a genome-wide significant burden of rare, damaging mutations in Ephrin B4 (EphB4).^32,33^ Interestingly, deletion of EphB4 in zebrafish recapitulates anomalies in the dorsal cranial vessels representing the phylogenic homologue of the vein of Galen.^33^ EphB4’s role as a critical regulator of AV specification^32,33^ suggests VOGM may share a common pathogenetic mechanism with other congenital AV shunts such as AV malformation (AVM). Specifically, incomplete AV differentiation and zonation are hallmarks of AVM, driven in part by EphB4^34^ mutations as well as several other genes regulating the developmental cerebrovascular niche.^8,35^ ETS, for the first time, enables *in situ* analysis to delineate these pathogenic mechanisms and pathways responsible for VOGM and cerebrovascular development more broadly.

The genetics underlying VOGM, and their relative contribution (compared to environmental factors) to disease, however, remain poorly understood.^30^ ETS represents a novel method for investigating these unanswered questions by screening lesional VOGM EC’s for *de novo*, somatic, and second hit mutations.^36,37^ Complementary *in situ* scRNA seq may elucidate vascular maintenance and pro-growth molecular programs that are not strictly genetic or heritable, but triggered in response to environmental factors. The PI3K/AKT/MTOR^38^ and RAS/RAF/MEK/ERK^39^ signaling programs, for example, influence angiogenesis, cell growth, apoptosis, and differentiation. Both are dysregulated in AVM, but at various frequency and intensity. Upstream EphB4 mutations are responsible in some instances yet absent in others. Additional mutations and transcriptional aberrations have been described in other vascular malformations, the vast majority being mosaic and restricted to pathologic tissue. This highlights the potential role for ETS and other evolving approaches such as liquid biopsy as tools to understand VOGM biology and to guide treatment.^40-43^

Future studies of ETS for genomic analysis may inform personalized therapies for VOGM and cerebrovascular diseases more broadly. While several existing compounds modulate RAS/RAF/MAPK and PI3K/AKT/mTOR ^44-47^ signaling, these treatments may be misguided without patient-specific data. Somatic WES and scRNA seq have the potential to identify disease-causing mutations and disease-propagating transcriptional aberrations. Both represent potential targets for gene therapy, novel or repurposed pharmacotherapies. However, protocols for reliably sequencing picogram DNA quantities and interpreting the “big data” inherent to scRNA seq remain a focus of ongoing study.

### Limitations

VOGM is a rare disease and treatment limited to few comprehensive pediatric hospitals with the requisite expertise. However, even at tertiary referral centers only 2-3 new VOGM cases may be seen annually. The small sample size reported here reflects this limitation, which reduces statistical power and challenges the generalizability of our results. We have formed the VOGM Genetics Research Consortium (VOGM-GRC, www.vogm-genetics.com), a multi-institutional consortium of leading pediatric centers across the world to address this shortcoming in future studies.

## Conclusions

We describe here a method for successfully performing ETS in VOGM that addresses unique properties of this formidable arteriovenous shunt. The potential benefits of ETS in VOGM are numerous and include (i) acquisition of lesional tissue to identify somatic mutations, (ii) ability to study molecular alterations in the shunt over time as they remodel, and (iii) enabling precision-medicine based approaches to pharmacologically targetable mutations. Nevertheless, ETS devices, sequencing platforms and genomic informatics all demonstrate substantial opportunities to improve. Our understanding of VOGM pathophysiology and vascular maintenance will grow in concert with such advances, potentially informing novel treatments to better outcomes across this specific population and cerebrovascular disease more broadly.

## Data Availability

All data produced in the present study are available upon reasonable request to the authors.

## Notes

**Conflict of Interest:** A.T.H. and J.G.J. received material support (i.e. coils) from Balt for the purposes of this study. Balt played no role in designing, collecting, interpreting, or reporting the data presented in this manuscript.

### Competing Interest Statement

Conflict of Interest: A.T.H. and J.G.J. received material support (i.e. coils) from Balt for the purposes of this study. Balt played no role in designing, collecting, interpreting, or reporting the data presented in this manuscript.
This paper has been submitted for presentation consideration at the American Association of Neurological Surgeons (AANS) and Congress of Neurological Surgeons (CNS) joint section for Pediatric Neurosurgery in Oklahoma City, OK (USA) in December 2023.

### Funding Statement

Disclosure of Funding: The Aneurysm and AVM Foundation (TAAF), Joe Niekro Foundation (JNF), Robert J. Dempsy, MD Research Award from the American Association of Neurological Surgeons (AANS) and Congress of Neurological Surgeons (CNS) joint section for Cerebrovascular Neurosurgery, Kaul Pediatric Research Institute (KPRI) of Childrens of Alabama, and the Center for Clinical and Translational Sciences (CCTS) at University of Alabama at Birmingham provided funding for this study. A.T.H. and Z.C. are supported by the National Institutes of Health (R21 NS135321). Z.C. also received support from the National Institutes of Health (R35 GM138212).

### Author Declarations

Ethics committee/IRB of University of Alabama at Birmingham gave ethical approval for this work.

### Summary of Updates

Updated text.

